# The burden of SARS-CoV-2 Infection and Severe Illness in South Africa March 2020-August 2022: A synthesis of epidemiological data

**DOI:** 10.1101/2024.10.30.24316404

**Authors:** Larisse Bolton, Stefano Tempia, Sibongile Walaza, Waasila Jassat, Kaiyuan Sun, Debbie Bradshaw, Rob Dorrington, Jackie Kleynhans, Neil Martenson, Anne von Gottberg, Nicole Wolter, Juliet R.C. Pulliam, Cheryl Cohen

**Affiliations:** South African Centre for Epidemiological Modelling and Analysis (SACEMA), School for Data Science and Computational Thinking, Stellenbosch University, Stellenbosch, South Africa; Centre for Respiratory Diseases and Meningitis, National Institute for Communicable Diseases of the National Health Laboratory Service, Johannesburg South Africa; School of Public Health, Faculty of Health Sciences, University of the Witwatersrand, Johannesburg, South Africa; Division of International Epidemiology and Population Studies, Fogarty International Center, National Institutes of Health, Bethesda, Maryland, United States of America; South Africa Medical Research Council, Burden of Disease Unit, Cape Town, South Africa; Department of Family Medicine and Public Health, University of Cape Town, South Africa; Centre for Actuarial Research (CARe), Faculty of Commerce, University of Cape Town; Perinatal HIV Research Unit, University of the Witwatersrand, Johannesburg, South Africa and Johns Hopkins University Center for TB Research; School of Pathology, Faculty of Health Sciences, University of the Witwatersrand, Johannesburg, South Africa

**Keywords:** SARS-CoV-2, national burden of disease, disease severity, public healthcare, disease pyramid

## Abstract

**Introduction:** Data on burden of SARS-CoV-2 infections by age group and for different severity levels are lacking. We estimated the South African SARS-CoV-2 disease burden and severity, describing changes in the shape of the disease burden pyramid with successive waves.

**Methods:** We estimated SARS-CoV-2 medically and non-medically attended illness stratified by severity (mild, severe non-fatal, and fatal) during the initial five waves, spanning 1 March 2020 through 13 August 2022. We utilised individual-level national surveillance, healthcare-utilisation and serosurvey data to calculate wave-specific hospitalisation-fatality (HFR) and infection-fatality ratios (IFR). We estimated wave-specific incidence rates per 100,000 population with 95% confidence intervals derived from bootstrapping the individual-level data.

**Results:** On 13 August 2022, the estimated cumulative number of SARS-CoV-2 infections in South Africa was 105 million, of which 399,886 (0.38%) were severe non-fatal and 258,754 (0.25%) fatal. 29% of severe non-fatal illness and 55% of deaths occurred outside hospital. The highest burden of severe and fatal illness was during the Delta wave (wave 3), and the HFR across the initial three waves was similar (range 31%-34%). Although there were more infections during the Omicron BA.1 wave (wave 4), there was a substantial reduction in HFR (14%). Successive waves saw a reduction in the rate of increase in mortality and hospitalisations with increasing age.

**Conclusions:** The substantial South African national burden of SARS-CoV-2 for the initial five waves contradicts the belief of minimal impact in Africa. A high proportion of severe non-fatal and fatal illness occurred outside of hospital, highlighting the importance of studies of health-seeking and vital registration systems to document the full burden of illness. Highest burden of severe illness and death was in the Delta wave. Following Omicron emergence severe illness reduced, and age-distribution for the incidence of medically attended severe non-fatal illness shifted to a J-shape, possibly reflecting the shift from widespread transmission to an endemic pattern.

**KEY MESSAGES:** *What is already known on this topic?:* As SARS-CoV-2 spread globally, initial assumptions painted a bleak picture for Africa due to its existing challenges in healthcare service delivery, multimorbidity, poverty and lack of resources needed to fight the infection. The number of cases and deaths reported during the pandemic seemed to contradict these initial assumptions. South Africa recorded over 4 million laboratory-confirmed cases of COVID-19 during the first three years of the pandemic. However, it is estimated that only a tenth of the cases were diagnosed. With the lack of testing, inconsistent healthcare-seeking behaviour, changes in attack, reinfection and symptomatic rates, the true burden of SARS-CoV-2 across the different age groups and severity levels were largely unknown. Although real-time epidemiological data was crucial for informing intervention strategies throughout the pandemic, it is now essential to quantify and describe the evolution of the epidemiology over successive pandemic waves as more information was made available.

*What this study adds?:* We found a high burden of severe illness and death in the first three waves of SARS-CoV-2, peaking in the third (Delta) wave. The emergence of the Omicron BA.1 variant was associated with very high rates of infection but substantial reductions in disease severity. Incidence of severe illness and hospitalisation fatality, generally increased with increasing age. Successive waves saw a reduction in the rate of the increase in mortality with increasing age and increases in hospitalisation fatality ratios in children below 5 years of age suggesting shift from the epidemic state to a J-shaped distribution in mortality, typical of seasonal respiratory viruses. Notably a high proportion of severe illness (29%) and death (55%) occurred outside hospital.

*How this study might affect research, practice or policy?:* Our study provides insights into the changes in patterns of infection and disease following introduction of a novel pathogen into a susceptible population which may be useful for future pandemic planning. The high proportion of undiagnosed and unreported illness and the high proportion of severe illness and death occurring outside of the hospital suggest that strengthening of access to diagnosis and care is needed in our setting.

## INTRODUCTION

As of the end of August 2023, confirmed numbers of COVID-19 cases and deaths worldwide exceeded 770 million and 6.9 million, respectively.^1^ South Africa is an upper middle-income country in Sub-Saharan Africa with a population of more than 60 million individuals.^2,3^ In 2022, 28% of the population was aged below 15 years and the population HIV prevalence was 12.6%.^3^ More than 4 million laboratory-confirmed cases of COVID-19 were identified in South Africa from the first confirmed case in early March 2020 until 25 March 2023^4^, cumulatively comprising almost half (42.7%) the total confirmed COVID-19 cases in Africa.^5^ SARS-CoV-2 testing capacity in Africa is limited and in South Africa, less than 10% of all SARS-CoV-2 cases during the first three waves are estimated to have been diagnosed.^6,7^ South Africa’s vaccination roll-out was initiated among healthcare workers in February 2021 and extended to the elderly and adolescents in May and October 2021, respectively.^8^ Vaccination coverage within South Africa reached 27% by 12 January 2022.^9^

From 5 March 2020 through 31 August 2022, South Africa experienced five waves of SARS-CoV-2, each dominated by a different variant (D614G, Beta, Delta, Omicron BA.1/2 and Omicron BA.4/5 respectively)^10^. As the virus becomes endemic with successive waves of infection, a rise in population SARS-CoV-2 immunity induced by natural infection and vaccination can lead to changes in the epidemiology of disease.^11^ These changes may include reductions in disease severity, as well as changes in the overall burden and age distributions of infections, illnesses and deaths. Furthermore, different emerging SARS-CoV-2 variants may be associated with immune evasion and variation in virulence, infectiousness and/or transmissibility, which can affect rates of infection and reinfection as well as variant’s intrinsic severity of illness.^12–14^

South Africa’s government funded^15^ public healthcare sector is the primary provider of healthcare for the majority of the population^16^, yet resources are limited^17^. Although public clinics, hospitals, or other institutions are the primary point of healthcare access for 72% of the population, healthcare-seeking behaviour varies.^16^ Considering the lack of testing as well as variable healthcare-seeking behaviour and changes in the symptomatic proportion, attack rates, and reinfection frequency across the five SARS-CoV-2 waves, the shape of the disease burden pyramid is largely unknown.

Data on the burden of SARS-CoV-2 in successive waves across different levels of severity are limited globally. This is particularly important in Africa, where resources are limited and official statistics suggest that the burden of cases and mortality was substantially lower than in other regions.^5^ We aimed to estimate the burden of SARS-CoV-2 infection and severe disease in South Africa from March 2020 through August 2022, and to describe the variations in the shape of the disease burden pyramid across the first five pandemic waves. At the start of the pandemic, it was critical to have realistic epidemiological data to model the impact of interventions. The models incorporated all accessible SARS-CoV-2 epidemiological data and were support tools to South African policy makers; updating stakeholders and the South African public throughout the course of the pandemic.^18^ However, now it is important to develop best estimates of the epidemiology, and how it changed over time as information about the spread of a novel respiratory infection became available.

## MATERIALS AND METHODS

### 1. Conceptual outline of burden strata with data sources

#### 1.1 Strata definitions

The SARS-CoV-2 burden comprises both asymptomatic infection (no clinical manifestation of disease) and symptomatic illness. We estimated the burden in three mutually exclusive severity strata: non-severe infection (asymptomatic infection and mild or moderate illness), severe non-fatal illness, and fatal illness.^19^ Severe non-fatal and fatal illness were further stratified into medically and non-medically attended illness (Figure 1). Individuals with severe illness who were admitted to hospital or individuals who died in hospital were considered medically attended. Individuals with severe illness who were not hospitalised and those who died out of hospital were considered non-medically attended.^19^

**Figure 1.**
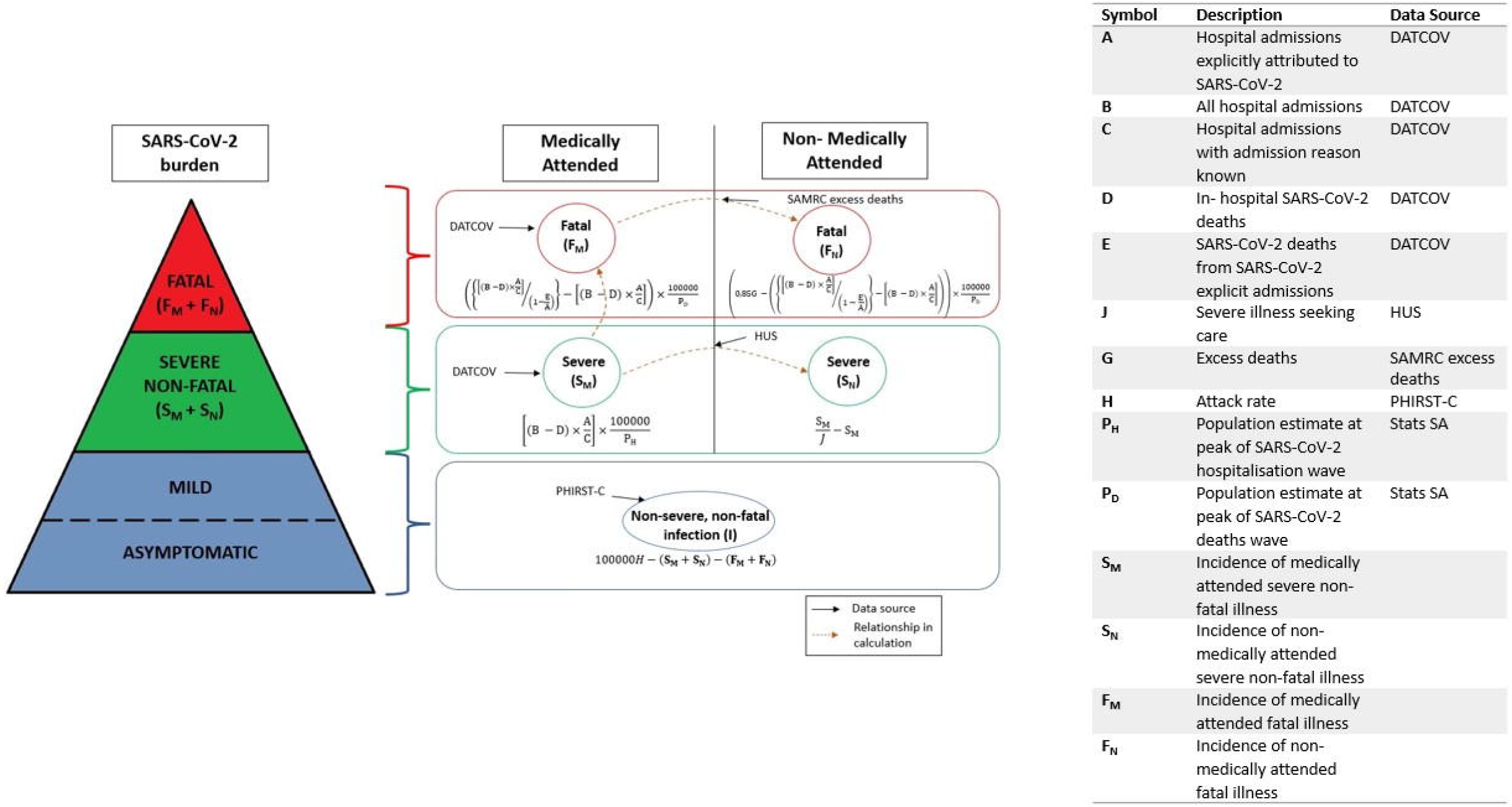
A summary illustration of the burden pyramid and related disease severity strata by medical attendance with data inputs and relationships between strata. (DATCOV = National surveillance programme for SARS-CoV-2 hospitalisations8, HUS = Healthcare utilization survey22, SAMRC excess deaths = South African Medical Research Council (SAMRC) estimates of excess deaths20, PHIRST-C = Prospective Household study of SARS-CoV-2, Influenza and Respiratory Syncytial virus community burden, Transmission dynamics and viral interaction in South Africa – Coronavirus disease 201923. For details on calculations, refer to Section 3.2.

#### 1.2 Data sources

1. *Individual-level SARS-CoV-2 public and private hospital admissions and subsequent in-hospital deaths from the DATCOV national surveillance programme for SARS-CoV-2 hospitalisations*.^8^ DATCOV includes data on COVID-19 hospitalisations from all private and public healthcare facilities in South Africa. Hospitalised patients comprised individuals with a positive SARS-CoV-2 test, irrespective of whether hospitalisation was attributable to SARS-CoV-2 or not. Reporting of all laboratory-confirmed cases was legally mandated. DATCOV variables included demographic details such as age and sex, admission information such as the date and reason for admission, and patient outcome such as death or discharge.
2. *South African Medical Research Council (SAMRC) estimates of excess deaths by sex and 5-year age bands*.^20^ The SAMRC, in partnership with the University of Cape Town (UCT), adapted the existing annual Rapid Mortality Surveillance (RMS) process to produce a near real-time (weekly) system for following and observing COVID-19 excess deaths. Weekly updates to the deaths recorded on the National Population Register (NPR), together with the classification of deaths due to natural or unnatural cause were obtained and adjusted for both under and late registration of deaths. Excess SARS-CoV-2 associated deaths are estimated by comparing observed to expected deaths derived from a negative binomial regression model of deaths in pre-COVID-19 years, described in detail elsewhere.^21^ While we attempted to account for the proportion of deaths attributable to SARS-CoV-2, robust data on this are lacking and it is possible that this varied over the pandemic.
3. *Healthcare utilisation surveys (HUS) conducted in three communities in three provinces*.^22^ From November 2020 through April 2021, fieldworkers enrolled 23,003 individuals from 5,804 randomly selected households. All individuals reporting severe respiratory illness (SRI) since the start of the SARS-CoV-2 pandemic were asked about healthcare-seeking behaviour including the proportion of these seeking care at healthcare facilities.
4. *Age-stratified infection attack rates by SARS-CoV-2 wave obtained from the Prospective Household study of SARS-CoV-2, Influenza and Respiratory Syncytial virus community burden, Transmission dynamics and viral interaction in South Africa – Coronavirus disease 2019 (PHIRST-C) cohort*.^23^ This cohort comprised 1,200 participants randomly selected from two communities in two provinces, including 643 participants from a rural site in Agincourt (Mpumalanga Province, South Africa) and 557 participants from an urban site in Jouberton (North-West Province, South Africa). Ten consecutive serum specimens were obtained from each participant between July 2020 and April 2022 and tested for anti-SARS-CoV-2 antibodies.^6,7,12,23^ The estimated age-stratified infection attack rates from the urban site were employed in calculations, because these were thought to be more representative of South Africa as a whole since the majority of South Africans live in urban areas.^24^ The infection attack rate was defined as the number of new infections divided by the population at risk over a defined period.^25^
5. *Mid-year population estimates*, unstratified and stratified in 5-year age bands and sex strata, provided by the government agency, Statistics South Africa.^26,27^

### 2. Definitions

#### 2.1 Wave

The total number of SARS-CoV-2 hospitalisations and in-hospital deaths were calculated by epidemiological year (epiyear) and epidemiological week (epiweek). The peaks (maximum SARS-CoV-2 hospitalisations and in-hospital deaths), as well as the nadirs (minimum hospitalisations and in-hospital deaths between these peaks) were then determined. For both hospitalisations and in-hospital deaths, the first wave was defined to extend from the start of the epiweek associated with the start of the pandemic in South Africa to the end of the epiweek of the nadir after this peak. The subsequent waves were defined to extend from the start of the epiweek following the nadir prior to the peak till the end of the epiweek of the nadir following the peak (Figure 2, Supplementary Table 1). However, to achieve agreement between the in-hospital and SAMRC excess deaths wave definitions, an epiweek was included at the end of wave 2 and the start of wave 3. Wave 5 was truncated at epiweek 32 of 2022. The SARS-CoV-2 hospitalisations and in-hospital deaths wave definitions were employed for the respective hospitalisation and in-hospital fatalities stratum estimates.

**Table 1.** Wave-specific incidence of severe non-fatal and fatal SARS-CoV-2 infections in South Africa by medical attendance per 100 000 population at risk from 01 March 2020 through 13 August 2022.

| Wave number/<br>Dominant variant | Medically attended |  | Non-medically attended |  | Total |  |  |
| --- | --- | --- | --- | --- | --- | --- | --- |
|  | *Incidence/100 000 population*<br>*(95% CI)* |  | *Incidence/100 000 population*<br>*(95% CI)* |  | *Incidence/100 000 population*<br>*(95% CI)* |  |  |
|  | Severe non-fatal | Fatal | Severe non-fatal | Fatal | Severe non-fatal | Fatal | Non severe non-fatal and non-fatal<sup>a</sup> |
| **1/**<br>**D614G** | 91.3<br>(90.7, 91.9) | 47.3<br>(45.6, 49.2) | 36.7<br>(24.5, 52.2) | 20.9 | 128.0<br>(116.1, 143.6) | 68.2 | 25 215.1<br>(18 421.8, 27 884.6) |
| **2/**<br>**Beta** | 129.1<br>(128.6, 129.7) | 61.3<br>(59.9, 62.8) | 51.9<br>(34.4, 73.8) | 81.7 | 181.0<br>(164.0, 202.8) | 143.0 | 23 829.5<br>(20 516.9, 29 106.6) |
| **3/**<br>**Delta** | 156.6<br>(155.9, 157.2) | 69.2<br>(67.6, 70.8) | 62.9<br>(41.8, 89.6) | 87.1 | 219.5<br>(199.2, 246.3) | 156.2 | 33 991.5<br>(28 477.3, 37 441.5) |
| **4/**<br>**Omicron BA.1/2** | 72.7<br>(72.2, 73.2) | 12.2<br>(11.8, 12.7) | 29.2<br>(19.4, 41.6) | 27.3 | 101.9<br>(92.3, 114.3) | 39.5 | 56 612.3<br>(53 114.7, 62 679.4) |
| **5/**<br>**Omicron BA.4/5** | 26.7<br>(26.3, 27.0) | 4.2<br>(3.9, 4.5) | 10.7<br>(7.1, 15.3) | 20.2 | 37.4<br>(34.0, 42.0) | 24.4 | 33 324.7<br>(31 033.6, 40 980.1) |

<sup>a</sup>Non-severe non-fatal and non-fatal = (Infection attack rate x 100 000) – Total incidence of severe non-fatal – Total incidence of fatal

**Figure 2.**
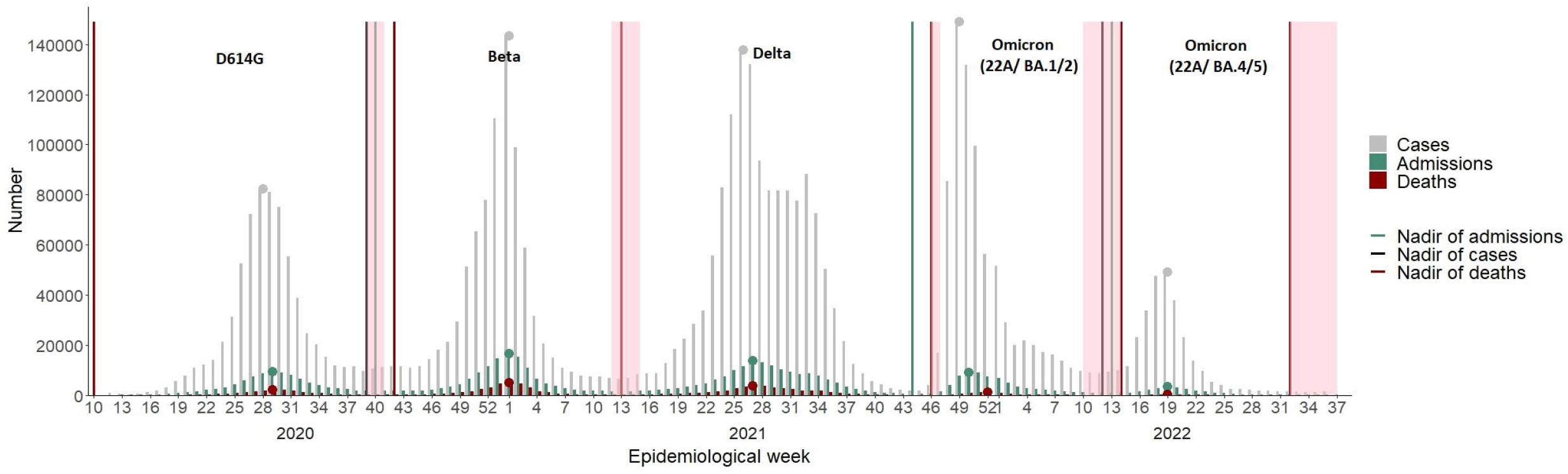
Number of confirmed SARS-CoV-2 cases4, hospital admissions and in-hospital deaths8 in South Africa 01 March 2020 through 13 August 2022 by epidemiological week as reported by the South African National Institute for Communicable Diseases (NICD). Black, green, and red lines define cases, hospitalisations and in-hospital death wave nadirs and grey, green, and red points define the peaks in cases, hospitalisations, and in-hospital deaths, respectively. Pink bands represent the time of blood draws for PHIRST-C. Dominant SARS-CoV-2 variant specified above each wave.23

#### 2.2 Country population at wave peak

The extracted mid-year population estimates (Section 1.2(e)) were obtained for each year, and the proportion weekly change in the population was estimated by dividing the proportion change in the population numbers between the epidemiological years and the number of weeks. Under the assumption of a linear weekly proportional change, the weekly population numbers were then interpolated by addition of the starting population and the calculated change.

### 3. Estimation approach

All analyses and calculations were carried out in RStudio 2023.06.0+421 for Windows^28^, running R Statistical software version 4.2.3^29^. The burden estimation approach is outlined hereafter; calculations for each stratum were implemented on an age-stratified and per-wave basis unless otherwise noted. Furthermore, national burden estimates were subsequently obtained from age-stratified figures. Figure 1 depicts the burden pyramid and related disease severity strata with data inputs and relationships between strata.

#### 3.1 Severe non-fatal and fatal illness

##### a) Medically attended severe non-fatal illness (incidence of hospitalisation)

Using the SARS-CoV-2 hospitalisation wave definitions, all hospital admissions (B), hospital admissions with admission reasons known (C), and hospital admissions explicitly attributed to SARS-CoV-2 (A) were extracted from DATCOV, respectively. The proportion of hospital admissions explicitly attributed to SARS-CoV-2 among all admissions with known admission reasons was defined as:

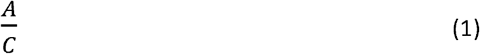

The total in-hospital deaths (D) were determined from DATCOV using the SARS-CoV-2 in-hospital deaths definitions. The adjusted number of severe, medically attended SARS-CoV-2 hospitalisations was obtained by subtracting the in-hospital deaths (D) from the total number of admissions (B) and multiplying the outcome by the proportion of explicitly attributed SARS-CoV-2 admissions (from (1)). That is,

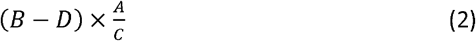

Finally, the medically attended severe non-fatal illness incidence rate per 100,000 (*SM*) was calculated by dividing the adjusted number of SARS-CoV-2 hospitalisations (from (2)) by the respective national population estimate at the peak of the SARS-CoV-2 hospitalisations wave (P_H_) and multiplying by 100,000:

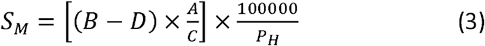

##### b) Medically attended fatal illness (incidence of in-hospital death)

The in-hospital explicitly attributed SARS-CoV-2 deaths from explicitly attributed SARS-CoV-2 hospital admissions (E) were determined from DATCOV using the SARS-CoV-2 in-hospital deaths wave definitions. The hospitalisation-fatality ratio (HFR) (defined as the proportion of SARS-CoV-2 deaths among SARS-CoV-2 hospitalised individuals)^30^ was determined as follows:

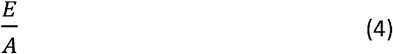

The adjusted number of total hospitalisations was then calculated by dividing the adjusted number of SARS-CoV-2 hospitalisations (from (2)) by the proportion of hospitalisations that did not result in death, i.e., 1 – HFR (from (4)). That is,

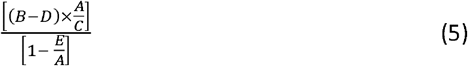

The medically attended fatal illness incidence rate per 100,000 (*F*_*M*_) was then determined by subtracting from (5) the adjusted SARS-CoV-2 hospitalisations (from (2)), dividing by the respective national population estimate at the peak of the SARS-CoV-2 in-hospital deaths wave (*P*_*D*_), and multiplying by 100,000:

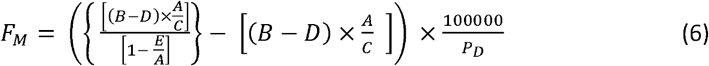

##### c) Non-medically attended severe non-fatal illness

The incidence of non-medically attended severe non-fatal illness (*S*_*N*_) is defined as the difference between the incidence of hospitalisation (*S*_*M*_) (from (3)) divided by the average proportion of severe illness seeking care from the HUS (J) and the incidence of hospitalisation (*S*_*M*_), i.e.,

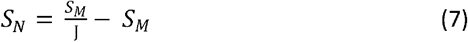

We assumed the average proportion of severe illness seeking care from the HUS remains unchanged irrespective of age strata or wave as the survey did not have sufficient power to stratify, and estimates were not available by wave.

##### d) Non-medically attended fatal illness (incidence of out-of-hospital deaths)

We assumed that 85% of excess deaths (G) were attributable to SARS-CoV-2 based on published data. The incidence of out-of-hospital deaths per 100,000 (*F*_*N*_) was then (8) calculated as the SARS-CoV-2 attributable excess deaths (0.85*G*) less the adjusted SARS-CoV-2 in-hospital deaths, divided by the respective national population estimates at the peak of the SARS-CoV-2 deaths waves (*P*_*D*_) times 100,000. That is,

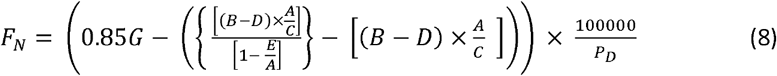

For the age-specific estimates, age groups for which in-hospital deaths exceeded the attributable excess deaths, the non-medically attended fatal illness estimate was set to zero, i.e., the SARS-CoV-2 attributable excess deaths were assumed equal to the in-hospital deaths.

#### 3.2 Non-severe, non-fatal illness (i.e., moderate, mild and asymptomatic illness)

The non-severe, non-fatal infection incidence (I) was obtained by multiplying the wave and age-specific attack rates (H) from PHIRST-C (refer to Section 1.2 (d)) by 100,000 and subtracting out the medically and non-medically attended severe non-fatal and fatal illness (from (3),(6) – (8)) since disease severity strata are assumed mutually exclusive:

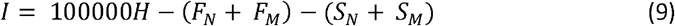

### 4. Estimation of uncertainty

Confidence intervals surrounding the incidence estimates were obtained from bootstrapping the individual-level data. That is, the wave-specific admission and in-hospital death data from DATCOV were each randomly sampled with replacement 1000 times stratified by wave. The average proportion of severe illness seeking care from the HUS was randomly generated from the binomial distribution with a probability of success out of a number of trails defined as the total number of individuals reporting severe illness across HUS sites given by the actual average and subsequently used within calculations for each of the 1000 replications on a wave-specific basis. It was assumed that the health seeking behaviour was consistent across age groups. Age-specific attack rates were obtained by simulating 1000 rates from the random binomial distribution with a probability of success defined as the actual attack rate from PHIRST-C and enforcing a threshold criterion to prevent the subsequent infection incidence from being negative. This threshold was defined to be the maximum of the sum of the 95% confidence interval upper bounds of the total incidence of severe non-fatal illness and total incidence of fatal illness. Should the generated random attack rate times 100 000 not be greater than the threshold, we continue to draw random attack rates until the threshold is exceeded. 95% confidence intervals were estimated as the 2.5 and 97.5 % quantiles of the respective bootstrapped/simulated estimates. No confidence intervals were generated for the incidence of out-of-hospital deaths and total incidence of death due to unavailability of line lists for bootstrapping. Confidence intervals for the incidence of non-severe non-fatal and non-fatal illness, IFR and SRIFR are solely derived from the uncertainty around the infection attack rates and incidence of severe non-fatal non medically and medically attended illness.

### 5. Ethical considerations and data sharing

Excess death data were provided in aggregate form; therefore, individuals’ consent was not required. The Human Research Ethics Committee (Medical) at the University of the Witwatersrand (Johannesburg, South Africa) approved the DATCOV protocol as part of a national surveillance programme (M160667). Because COVID-19 is a notifiable disease, individual patient consent was waived. The PHIRST-C and HUS protocol were approved by the University of Witwatersrand, Johannesburg, South Africa, Human Research Ethics Committee (reference 150808 and M200862, respectively). The data for this analysis was obtained from several different sources. Requests for the underlying data should be made from the custodians of the data. Aggregated DATCOV data are available on request to the South African National Institute of Communicable Diseases (NICD). The data dictionary is available on request to the data custodian, Dr Waasila Jassat. The analysis code and details of data custodians are available in the Github repository located at https://github.com/BoltonL/Burden_estimation_public. Detailed computational outputs from the analyses are provided in the Supplementary materials.

### 6. Patient and Public Involvement

The study involved a synthesis of retrospective data and thus it was not possible to involve patients or the public in the design, conduct, reporting or dissemination plans of this research.

## RESULTS

### Overall

Overall, across the first five waves of SARS-CoV-2 in South Africa, from 1 March 2020 through 13 August 2022, we estimate that there were 104,577,430 SARS-CoV-2 infection episodes, of which 399,886 (0.38%) were severe non-fatal illness episodes and 258,754 (0.25%) deaths out of a population of approximately 300 million individuals under observation over the period (Figure 2). 29% of severe non-fatal illness and 55% of deaths occurred outside of hospital.

### Severe disease and death overall

Incidence of both medically- and non-medically attended severe non-fatal and fatal illness increased from wave 1 to 3, with highest incidence occurring during the Delta wave (wave 3) (Figure 3, Table 1). The incidence of death, both medically- and nonmedically attended, also increased across the first three waves, peaking during the Delta (3^rd^) wave, with 69.2 in-hospital and 87.1 out-of-hospital deaths per 100,000 population, respectively. Thereafter, severe illness and death incidences decreased progressively in waves 4 and 5. The hospitalisation-fatality ratio (HFR), was approximately 32% in the first 3 waves, decreasing slightly from wave one to three (from 34% to 31%), but then dropped substantially in wave four and five (to 14%). In contrast, the out-of-hospital severe respiratory infection-fatality ratio (SRIFR) showed no consistent trend ranging from 37% to 65% across all five waves (Table 2).

**Table 2.**
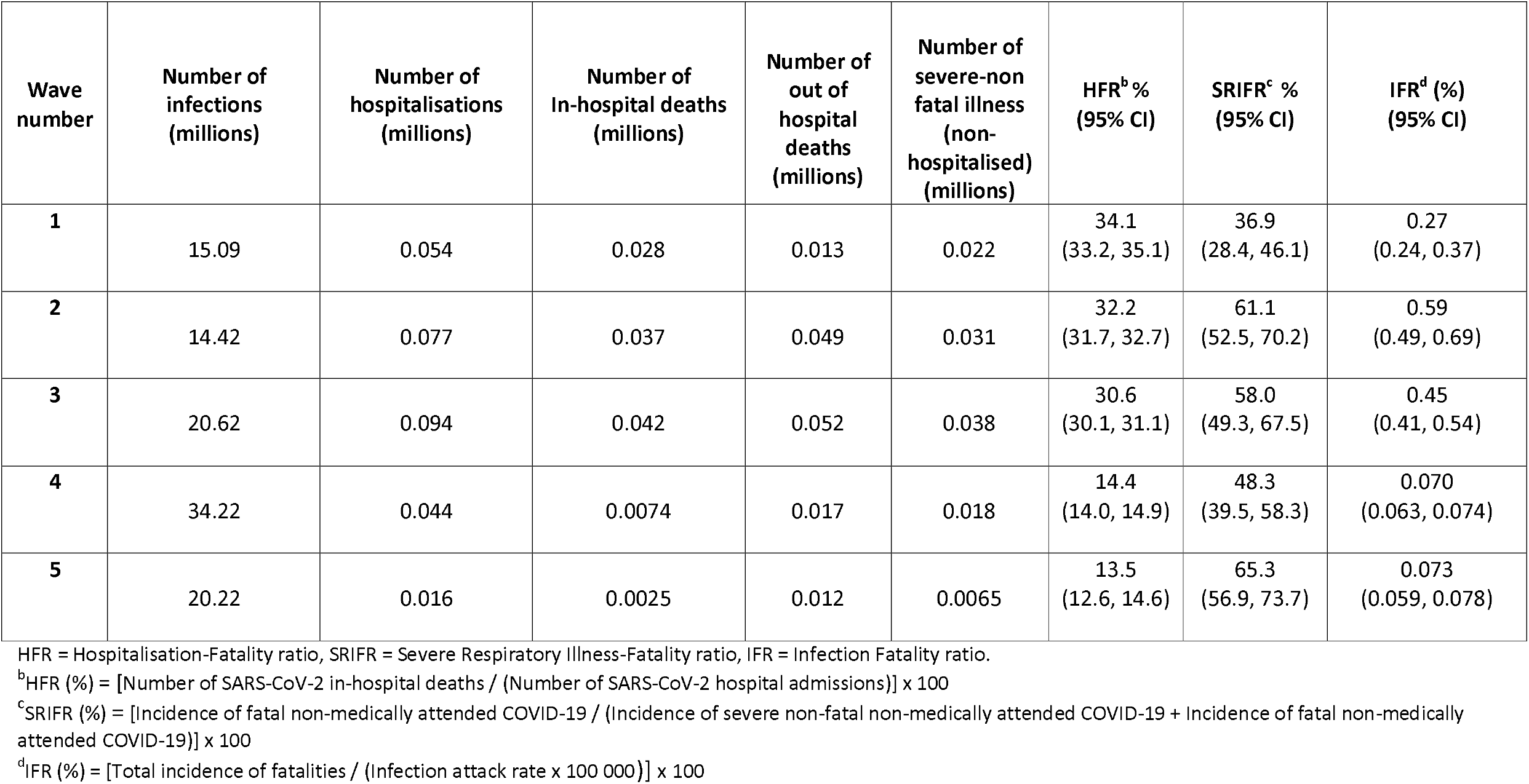
Wave-specific total numbers of SARS-CoV-2 infections, hospitalisations and deaths and hospitalisation-, severe respiratory illness – and infection-fatality ratio in South Africa from 01 March 2020 through 13 August 2022 (HFR = Hospitalisation-Fatality ratio, SRIFR = Severe Respiratory Illness-Fatality ratio, IFR = Infection-Fatality ratio).

**Figure 3.**
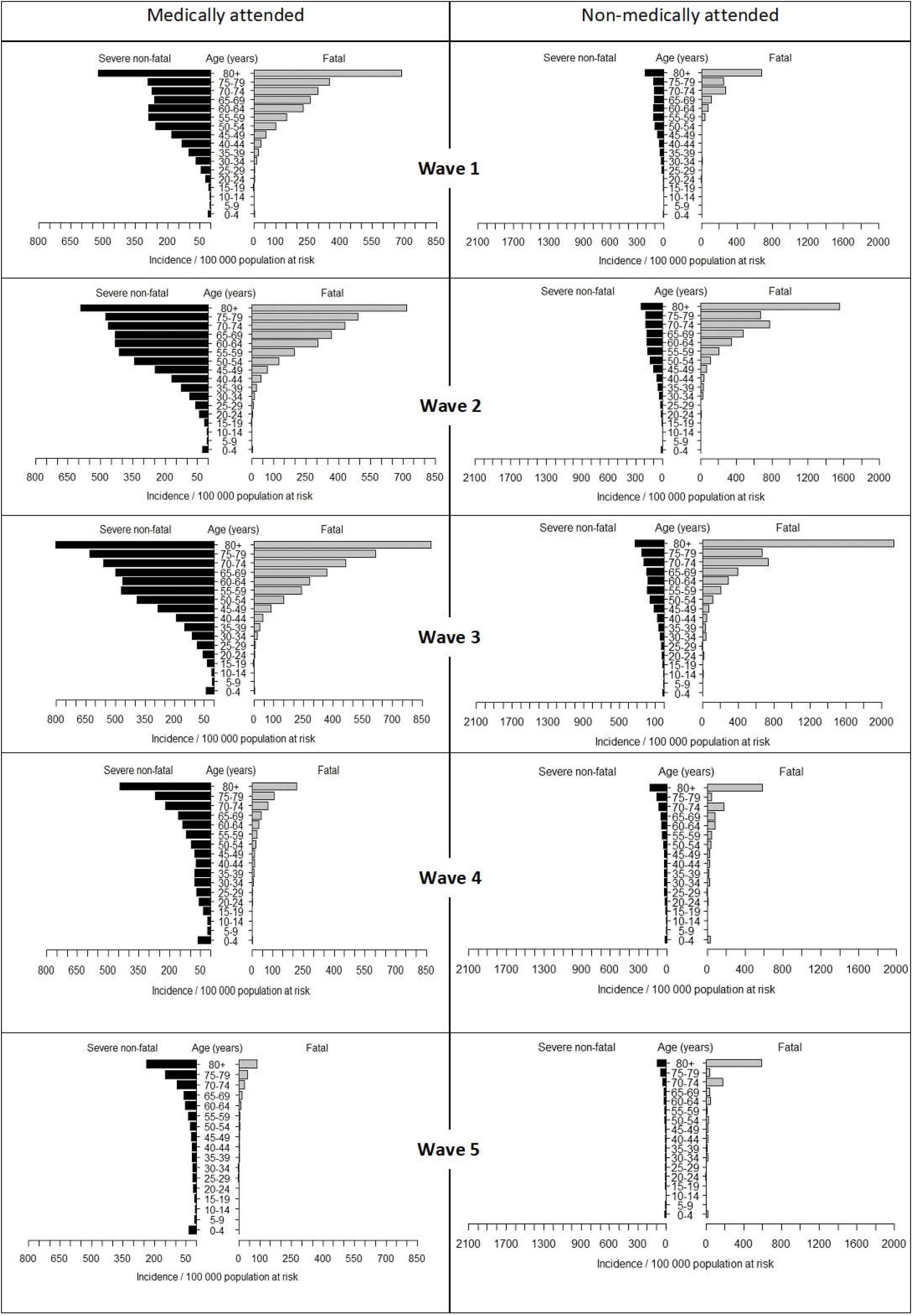
Age-specific incidence of severe and fatal illness by age for the first five SARS-CoV-2 waves in South Africa by medical attendance from 01 March 2020 through 13 August 2022.

### Infections and non-severe non-fatal illness overall

Similar to severe illness and death, the number of SARS-CoV-2 infections increased over the first three waves from 15.1 million in wave one to 20.6 million in wave three (Table 2). Following the emergence of the Omicron variant in wave 4 the number of infections peaked in wave 4 at 34.2 million and decreased thereafter in wave 5 to 20.2 million. In the first three waves the infection fatality ratio (IFR) ranged between 0.27%-0.59% and was highest in the second (Beta) wave. In the fourth and fifth waves the IFR dropped substantially to 0.07%.

### Severe disease and death by age group

The incidence of medically and non-medically attended severe non-fatal and fatal illness for those aged 65 years and older increased noticeably from wave 1 to wave 2 and a further sharp increase for individuals 75 years and older in wave 3 (Figure 3). The incidence of medically attended severe non-fatal and fatal illness in those aged 80 years and over was 802.5 (95%CI: 768.6-835.3) and 894.1 (95% CI: 815.7-978.2) per 100,000, respectively in wave 3. Following the emergence of the Omicron variant in wave 4, the incidence of severe non-fatal illness and deaths decreased markedly among individuals aged 15 years and older. Among individuals aged under 15 years incidence of severe illness was slightly increased in the fourth wave reducing again in the fifth wave. Deaths among individuals aged less than 15 years remained low in all five waves. Generally, as infection waves progress, the age-specific incidence of medically attended severe non-fatal illness approaches a J-shape curve (higher at the youngest and oldest ages, but higher at the oldest ages).

### Changes in HFR and IFR with age

Generally, the HFR increased with increasing age, showing a small increase in young children in some waves (Figure 4, Supplementary Figure 1). In wave 1, the 10-to-14-year-olds had the lowest HFR of 6% and those 80 years and older, the greatest at 57%. In individuals aged 30 years and older there was a marked drop in HFR in the fourth and fifth waves compared to the first three waves. Individuals aged 80 years and older had the highest HFR across waves and only experienced a 53% reduction in HFR ratio from wave one to five. In contrast, the HFR for individuals aged 30 years and older had a noticeably lower rate across age groups in waves 4 and 5 than waves 1 to 3. Consider for example, in wave 1 the difference in HFR between 30-to-34-year-olds and 80-years and above is about 42%, whilst in wave 5 this difference is around 18%. Like the HFR, the older age groups (65 years and above) experienced the highest IFR across all waves. For all waves the IFR was extremely low in individuals aged <40 years.

**Figure 4.**
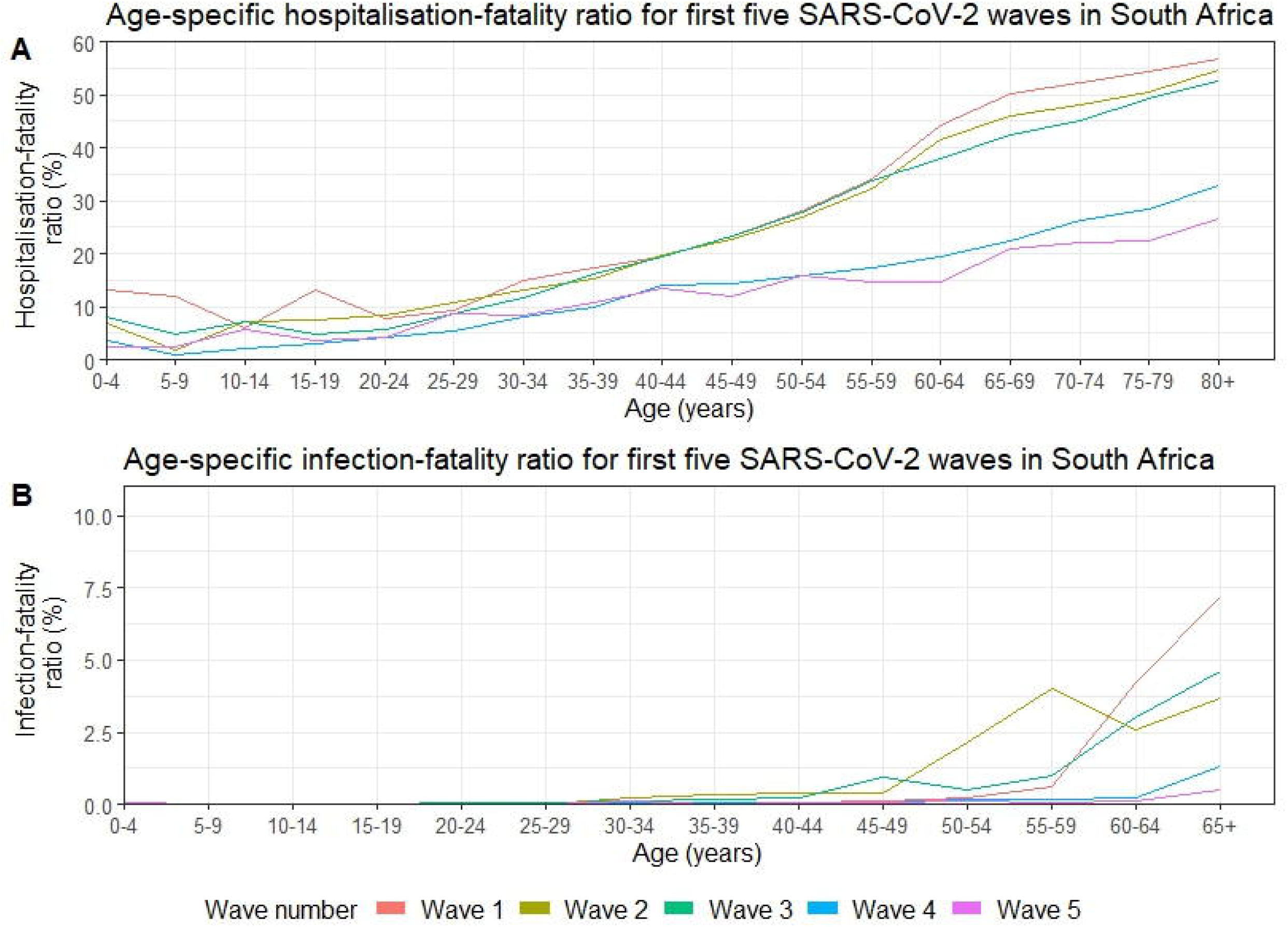
Age-specific fatality ratios (in %) for the first five SARS-CoV-2 waves in South Africa from 01 March 2020 through 13 August 2022. A. Hospitalisation-Fatality (%) B. Infection-Fatality (%).

## DISCUSSION

We estimate the burden of SARS-CoV-2 infection and disease severity in South Africa over the first five waves of infection to understand changes in disease burden with successive waves. We found a high burden of severe illness and death in the first three waves of SARS-CoV-2, peaking in the third (Delta) wave. The emergence of the Omicron BA.1 variant was associated with very high rates of infection but substantial reductions in disease severity. Incidence of severe illness and hospitalisation fatality, generally increased with increasing age. Successive waves saw a reduction in the rate of the increase in mortality with increasing age and increases in hospitalisation fatality ratios in children below 5 years of age suggesting shift from the epidemic state to a J-shaped distribution in mortality, typical of seasonal respiratory viruses. Notably a high proportion of severe illness (29%) and death (55%) occurred outside hospital.

The third (Delta) wave of infection had the highest incidence of severe non-fatal and fatal illness. Wave 4 (Omicron BA.1/2 variant) was characterised by marked reductions in severe disease; however, the incidence of asymptomatic, mild, or moderate disease was highest in this wave. Increasing severe disease in the Beta^14^ and Delta^32^ waves has been described from South Africa and other countries, possibly related to increased virulence and relaxation of restrictions in later waves. The high attack rates and lower severity following Omicron emergence can be explained in part by the increased transmissibility and immune evasive properties of Omicron.^12,33^ Together these led to very high infection attack rates. The high proportion of breakthrough infections in the Omicron wave likely contributed to reduced overall severity together with changes in virus tropism increasing the proportion of infections in the upper airway.^34^

Successive waves of infection were associated with shifts in the age distribution of severe disease. In all five waves, severe disease was concentrated in the elderly as has been described previously, driven in part by increasing comorbidities in this age group.^14,35,36^ Following the emergence of the Omicron variant, the reductions in severe disease were mostly seen in older adults, leading to a flattening of the steepness of increasing severity with increasing age and proportionately more severe disease in young children. This shift towards an age pattern more typical of seasonal respiratory viruses where severe disease is concentrated in the elderly and young children, could represent the transition from a pandemic to endemic epidemiology.^11^ Notably, despite the overall reduction in disease severity, more children and adolescents were hospitalised during the Omicron wave (BA.1/2 variant) than in the previous infection waves. This pattern of proportionately more disease in children following Omicron emergence in South Africa has been described previously, however not in the context of estimation of the burden pyramid in all age groups.^8,37,38^ Possible reasons for the shift in age distribution include changes in population immunity, pathogen virulence or changes in health seeking behaviour or testing practices. Persons aged 60 years and older were prioritised for vaccination in South Africa as from May 2021 and extended to 12 to 17 year olds in October 2021.^8^ This may also have contributed to the sharp decline in HFR among the elderly from wave 1 to wave 5 since vaccination coverage within South Africa reached 27% by 12 January 2022.^9^ However, the reduction in mortality in people living with HIV was not as dramatic with the emergence of the Omicron waves compared to HIV negative.^39^

A high proportion of severe disease and deaths occurred outside of hospitals^40^, highlighting the importance of studies of healthcare-seeking and access and strengthening of vital registration systems to document the full burden of illness.^20^ The high number of severe cases at the peaks of the waves caused pressure on the healthcare system, potentially reducing access and exacerbating underlying health inequities.^22,41^ An important limitation of our study is that we only had healthcare utilisation data from three communities in three provinces for the first two waves of the pandemic.^22,41^ Our assumption that healthcare seeking remained constant may not be valid and could be the reason why we do not see clear trends in the SRIFR over successive waves as we do for HFR.

Our study had other limitations. Out of hospital deaths were ascertained from modelling of excess COVID-19-associated deaths, using all cause vital registration data, thus not all excess deaths may have been due to COVID-19. We accounted for this by assuming that the attributable proportion was fixed for all waves; if the proportion did vary over time, this could have led to bias in estimates. Data for the different strata of severity were derived from different sources. For health seeking and infection attack rates, national data were not available, and we had to extrapolate nationally under the assumption that the available data were representative. If these measures varied by geographical region, this would have led to biased estimates. Although the infection attack rates accounted for reinfections, the estimated hospitalisations and deaths within our study occur as they do, whether they result from reinfections or not. Within our estimation of the medically attended severe non-fatal and fatal illness, it must be noted that the proportion of in-hospital deaths attributed to SARS-CoV-2 may not necessarily have been the same as the proportion of SARS-CoV-2 hospitalisation. Finally, we used the mid-year population estimates from 2020 through 2022 provided by Statistics South Africa as our population denominators throughout this study. These estimates are obtained from the Spectrum Policy Modelling system.^42^ However, the Thembisa model is also available to obtain population estimates.^3^ The total population estimated by the latter model (~59 million)^3^ is different from the recently published Census 2022 (~62 million)^43^ due to an underestimation of those 55 years and older.

In conclusion, we have demonstrated a high burden of infection and severe disease associated with SARS-CoV-2 over five successive waves in South Africa. With the onset of SARS-CoV-2 it was assumed that Africa would carry a large burden of infection and mortality due to weak health systems, lack of access to therapeutics and vaccines, poverty, malnutrition and high prevalence of underlying illness such as HIV and tuberculosis^44,45^, as well as high levels of persons left undiagnosed and untreated or not effectively treated. While reports of confirmed cases suggest that Africa was relatively spared, our analysis combining multiple data sources, suggests that this was not the case for South Africa. The estimated cumulative SARS-CoV-2 fatalities in South Africa across the first five waves (258,754) exceeds the number of confirmed SARS-CoV-2 deaths of high-income countries such as Germany (150 237) and the United Kingdom (205 780).^46^ Even more stark, our estimated cumulative SARS-CoV-2 cases over this period (105 million) far exceeds that reported for these high-income countries, namely 31.6 and 23.4 million, respectively.^46^ Furthermore, despite differences in methodology, our estimated IFR for wave 1 (0.27%) was comparable to that of Leticia, Columbia (0.28%).^47^ The discrepancy between low numbers of confirmed cases and actual number of infections were similarly observed across six districts in Zambia. During the first wave, a SARS-CoV-2 prevalence study reported 454 708 infections compared to a mere 4 917 laboratory confirmed cases.^48^ In future, it will be important to estimate the ongoing age-specific burden of COVID-19 compared to other respiratory viral illnesses in order to decide on priorities for prevention interventions such as vaccination.

## Data Availability

The data for this analysis was obtained from several different sources. Requests for the underlying data should be made from the custodians of the data. Aggregated DATCOV data are available on request to the South African National Institute of Communicable Diseases (NICD). The data dictionary is available on request to the data custodian, Dr Waasila Jassat.

https://github.com/BoltonL/Burden_estimation_public

## AUTHOR CONTRIBUTIONS

LB: formal analyses, investigation, software, validation, visualisation, writing – original, writing – review and editing. ST: conceptualisation, supervision, methodology, writing – subject-specific commentary in original, writing – review and editing. CC: conceptualisation, supervision, methodology, project administration, data provision, writing – subject-specific commentary in original, writing – review and editing, resources. JP: supervision, methodology, writing – review. SW: data collection, advisor, writing – review and editing. DB and RD: methodology, primary data – SAMRC excess deaths, writing – subject-specific commentary in original, writing – review and editing. WJ: primary data – DATCOV, writing - review and editing. KS: analyses of serological data, writing - review and editing. JK, NM, AvG: data collection – PHIRST-C, writing – review and editing. NW – data collection – PHIRST-C, HUS, writing – review and editing.

## ACKNOWLEDGMENTS

This work is based on research supported by the South African National Research Foundation (NRF). Any opinion, finding, and conclusion or recommendation expressed in this material is that of the authors and the NRF does not accept any liability in this regard. We acknowledge the support provided by OpenAI’s ChatGPT, which offered AI-driven insights that facilitated our editorial and review processes in the preparation of the manuscript by providing feedback and suggestions to improve the manuscript.

## FIGURE LEGENDS

**Supplementary figure 1.**
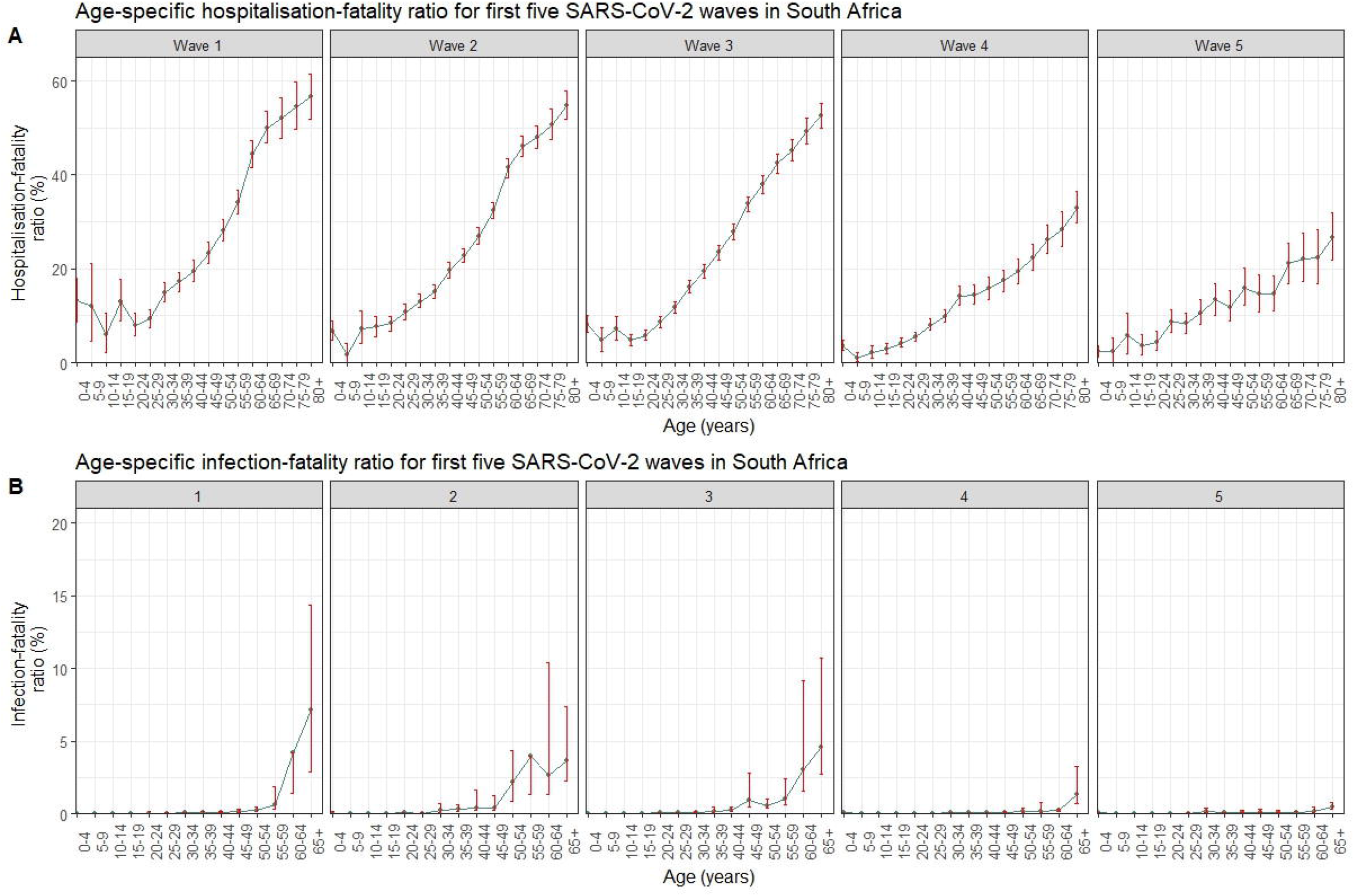
Age-specific fatality ratios (in %) with 95% confidence intervals for the first five SARS-CoV-2 waves in South Africa from 01 March 2020 through 13 August 2022. A. Hospitalisation-Fatality ratio (%) B. Infection-Fatality ratio (%).

## REFERENCES

1. Emergency Response, World Health Organisation (WHO). Weekly Epidemiological Update on COVID-19 - 1 September 2023. World Health Organisation (WHO); 2023. Accessed September 19, 2023. https://www.who.int/publications/m/item/weekly-epidemiological-update-on-covid-19 1-september-2023

2. The World Bank. World Bank list of economies. Published online 2020. Accessed June 26, 2023. https://datahelpdesk.worldbank.org/knowledgebase/articles/906519-world-bank-country-and-lending-groups

3. Johnson LF, Meyer-Rath G, Dorrington RE, et al. The Effect of HIV Programs in South Africa on National HIV Incidence Trends, 2000–2019. JAIDS Journal of Acquired Immune Deficiency Syndromes. 2022;90(2):115. doi:10.1097/QAI.0000000000002927

4. National Institute for Communicable Diseases (NICD). COVID-19 Weekly Epidemiology Brief: Week Ending 25 March 2023. National Institute for Communicable Diseases (NICD), South Africa; 2023. Accessed September 19, 2023. https://www.nicd.ac.za/diseases-a-z-index/disease-index-covid-19/surveillance-reports/weekly-epidemiological-brief/

5. World Health Organization (WHO). WHO Coronavirus (COVID-19) Dashboard. Published September 13, 2023. Accessed September 19, 2023. https://covid19.who.int/

6. Kleynhans J, Tempia S, Wolter N, et al. SARS-CoV-2 Seroprevalence in a Rural and Urban Household Cohort during First and Second Waves of Infections, South Africa, July 2020–March 2021. Emerg Infect Dis. 2021;27(12):3020–3029. doi:10.3201/eid2712.211465

7. Kleynhans J, Tempia S, Wolter N, et al. SARS-CoV-2 Seroprevalence after Third Wave of Infections, South Africa. Emerg Infect Dis. 2022;28(5):1055–1058. doi:10.3201/eid2805.220278

8. Jassat W, Abdool Karim SS, Mudara C, et al. Clinical severity of COVID-19 in patients admitted to hospital during the omicron wave in South Africa: a retrospective observational study. The Lancet Global Health. 2022;10(7):e961–e969. doi:10.1016/S2214-109X(22)00114-0

9. Suleman M, Lucero-Prisno Iii DE. South Africa’s COVID-19 vaccine rollout amid theemergence of Omicron. Popul Med. 2022;4(January):1–2. doi:10.18332/popmed/145772

10. National Institute for Communicable Diseases (NICD). SARS-CoV-2 Sequencing Update 24 March 2023. National Institute for Communicable Diseases (NICD), South Africa; 2023:28. Accessed January 16, 2024. https://www.nicd.ac.za/wp-content/uploads/2023/03/Update-of-SA-sequencing-data-from-GISAID-24-Mar-2023-final.pdf

11. Cohen C, Pulliam J. COVID-19 infection, reinfection, and the transition to endemicity. The Lancet. 2023;401(10379):798–800. doi:10.1016/S0140-6736(22)02634-4

12. Sun K, Tempia S, Kleynhans J, et al. Rapidly shifting immunologic landscape and severity of SARS-CoV-2 in the Omicron era in South Africa. Nat Commun. 2023;14(1):246. doi:10.1038/s41467-022-35652-0

13. Wolter N, Jassat W, Walaza S, et al. Early assessment of the clinical severity of the SARS-CoV-2 omicron variant in South Africa: a data linkage study. The Lancet. 2022;399(10323):437–446. doi:10.1016/S0140-6736(22)00017-4

14. Jassat W, Mudara C, Ozougwu L, et al. Difference in mortality among individuals admitted to hospital with COVID-19 during the first and second waves in South Africa: a cohort study. The Lancet Global Health. 2021;9(9):e1216–e1225. doi:10.1016/S2214-109X(21)00289-8

15. National Treasury: Republic of South Africa. Budget Review 2019. Published online February 20, 2019. Accessed April 17, 2023. https://www.treasury.gov.za

16. Statistics South Africa. General Household Survey 2021. Statistics South Africa; 2022. Accessed February 14, 2023. https://www.statssa.gov.za/

17. Marten R, McIntyre D, Travassos C, et al. An assessment of progress towards universal health coverage in Brazil, Russia, India, China, and South Africa (BRICS). The Lancet. 2014;384(9960):2164–2171. doi:10.1016/S0140-6736(14)60075-1

18. Meyer-Rath G, Hounsell RA, Pulliam JR, et al. The role of modelling and analytics in South African COVID-19 planning and budgeting. Shim E, ed. PLOS Glob Public Health. 2023;3(7):e0001063. doi:10.1371/journal.pgph.0001063

19. Tempia S, Walaza S, Moyes J, et al. Quantifying How Different Clinical Presentations, Levels of Severity, and Healthcare Attendance Shape the Burden of Influenza-associated Illness: A Modeling Study From South Africa. Clinical Infectious Diseases. 2019;69(6):1036–1048. doi:10.1093/cid/ciy1017

20. Dorrington RE, Moultrie TA, Laubscher R, Groenewald PJ, Bradshaw D. Rapid mortality surveillance using a national population register to monitor excess deaths during SARS-CoV-2 pandemic in South Africa. Genus. 2021;77(1):19. doi:10.1186/s41118-021-00134-6

21. Bradshaw D, Dorrington R, Laubscher R, Groenewald P, Moultrie T. COVID-19 and all-cause mortality in South Africa – the hidden deaths in the first four waves. S Afr J Sci. 2022;118(5/6). doi:10.17159/sajs.2022/13300

22. Wolter N, Tempia S, von Gottberg A, et al. Healthcare utilization during the first two waves of the COVID-19 epidemic in South Africa: a cross-sectional household survey.

23. Cohen C, Kleynhans J, Von Gottberg A, et al. SARS-CoV-2 incidence, transmission, and reinfection in a rural and an urban setting: results of the PHIRST-C cohort study, South Africa, 2020–21. The Lancet Infectious Diseases. 2022;22(6):821–834. doi:10.1016/S1473-3099(22)00069-X

24. Statistics South Africa. General Household Survey 2019. Statistics South Africa; 2019. https://doi.org/www.statssa.gov.za

25. Division of Reproductive Health, National Center for Chronic Disease Prevention and Health Promotion, US Centers for Disease Control and Prevention (CDC). Epidemiology Glossary. Reproductive Health. Published January 21, 2015. Accessed September 20, 2023. https://www.cdc.gov/reproductivehealth/data_stats/glossary.html

26. Statistics South Africa. Mid-Year Population Estimates 2022. Statistics South Africa; 2022:1–43. Accessed March 15, 2023. https://www.statssa.gov.za/?page_id=1854&PPN=P0302&SCH=73305

27. Statistics South Africa. Country-projection by population group, sex and age (2002-2022). Published online July 28, 2022. Accessed April 5, 2023. https://www.statssa.gov.za

28. RStudio Team. RStudio: Integrated development environment for R. Published online June 5, 2023. Accessed April 6, 2023. https://rstudio.com/

29. R Core Team. R: A language environment for statistical computing. Published online March 15, 2023. Accessed April 6, 2023. https://www.r-project.org

30. Epidemic and Pandemic Preparedness and Prevention (EPP), WHO Headquarters (HQ), WHO Worldwide. Estimating Mortality from COVID-19. World Health Organisation (WHO); 2020:1–4. Accessed September 19, 2023. https://www.who.int/publications/i/item/WHO-2019-nCoV-Sci-Brief-Mortality-2020.1

31. Moultrie T, Dorrington R, Laubscher R, Groenewald P, Bradshaw D. Correlation of Excess Natural Deaths with Other Measures of the COVID-19 Pandemic in South Africa. Burden of Disease Research Unit South African Medical Research Council; 2021:1–14. Accessed May 20, 2024. https://www.samrc.ac.za/sites/default/files/bod/weeklyreports/CorrelationExcessDeaths.pdf

32. Fisman DN, Tuite AR. Evaluation of the relative virulence of novel SARS-CoV-2 variants: a retrospective cohort study in Ontario, Canada. CMAJ. 2021;193(42):E1619–E1625. doi:10.1503/cmaj.211248

33. Garcia-Beltran WF, Lam EC, St. Denis K, et al. Multiple SARS-CoV-2 variants escape neutralization by vaccine-induced humoral immunity. Cell. 2021;184(9):2372–2383.e9. doi:10.1016/j.cell.2021.03.013

34. Meng B, Abdullahi A, Ferreira I, et al. Altered TMPRSS2 usage by SARS-CoV-2 Omicron impacts infectivity and fusogenicity. Nature. 2022;603(7902):706–714. doi:10.1038/s41586-022-04474-x

35. Shi Y, Yu X, Zhao H, Wang H, Zhao R, Sheng J. Host susceptibility to severe COVID-19 and establishment of a host risk score: findings of 487 cases outside Wuhan. Crit Care. 2020;24(1):108. doi:10.1186/s13054-020-2833-7

36. Cox MJ, Loman N, Bogaert D, O’Grady J. Co-infections: potentially lethal and unexplored in COVID-19. The Lancet Microbe. 2020;1(1):e11. doi:10.1016/S2666-5247(20)30009-4

37. Cloete J, Kruger A, Masha M, et al. Paediatric hospitalisations due to COVID-19 during the first SARS-CoV-2 omicron (B.1.1.529) variant wave in South Africa: a multicentre observational study. The Lancet Child & Adolescent Health. 2022;6(5):294–302. doi:10.1016/S2352-4642(22)00027-X

38. Chiwandire N, Jassat W, Groome M, et al. Changing Epidemiology of COVID-19 in Children and Adolescents Over Four Successive Epidemic Waves in South Africa, 2020–2022. Journal of the Pediatric Infectious Diseases Society. 2023;12(3):128–134. doi:10.1093/jpids/piad002

39. Jassat W, Mudara C, Ozougwu L, et al. Trends in COVID-19 admissions and deaths among people living with HIV in South Africa: analysis of national surveillance data. The Lancet HIV. 2024;11(2):e96–e105. doi:10.1016/S2352-3018(23)00266-7

40. Sabet N, Omar T, Milovanovic M, et al. Undiagnosed Pulmonary Tuberculosis (TB) and Coronavirus Disease 2019 (COVID-19) in Adults Dying at Home in a High-TB-Burden Setting, Before and During Pandemic COVID-19: An Autopsy Study. Clinical Infectious Diseases. 2023;77(3):453–459. doi:10.1093/cid/ciad212

41. Jassat W, Ozougwu L, Munshi S, et al. The intersection of age, sex, race and socio-economic status in COVID-19 hospital admissions and deaths in South Africa (with corrigendum). S Afr J Sci. 2022;118(5/6). doi:10.17159/sajs.2022/13323

42. Statistics South Africa. Mid-Year Population Estimates 2021. Department of Statistics; 2022. Accessed November 29, 2022. https://doi.org/www.statssa.gov.za

43. Statistics South Africa. Census 2022. Statistics South Africa; 2023:1–100. https://census.statssa.gov.za/assets/documents/2022/P03014_Census_2022_Statistical_Release.pdf

44. Senthilingam M. Covid-19: Why Africa’s pandemic is different. BMJ. Published online October 19, 2021:n2512. doi:10.1136/bmj.n2512

45. Katz MA, Schoub BD, Heraud JM, Breiman RF, Njenga MK, Widdowson MA. Influenza in Africa: Uncovering the Epidemiology of a Long-Overlooked Disease. Journal of Infectious Diseases. 2012;206(suppl 1):S1–S4. doi:10.1093/infdis/jis548

46. Global Change Data Lab, University of Oxford. COVID-19 Data Explorer. Our world in data. Published 2024. Accessed May 20, 2024. https://ourworldindata.org/explorers/coronavirus-data-explorer

47. Levin AT, Owusu-Boaitey N, Pugh S, et al. Assessing the burden of COVID-19 in developing countries: systematic review, meta-analysis and public policy implications. BMJ Glob Health. 2022;7(5):e008477. doi:10.1136/bmjgh-2022-008477

48. Mulenga LB, Hines JZ, Fwoloshi S, et al. Prevalence of SARS-CoV-2 in six districts in Zambia in July, 2020: a cross-sectional cluster sample survey. The Lancet Global Health. 2021;9(6):e773–e781. doi:10.1016/S2214-109X(21)00053-X

