## Supplementary document for "The burden of SARS-CoV-2 Infection and Severe Illness in South Africa March 2020-August 2022: A synthesis of epidemiological data"

**Supplement**

Supplementary table 1: Wave timing definitions for the first five SARS-CoV-2 waves in South Africa

| Wave number | Cases | | | Hospitalisations | | | Deaths | | |
|---|---|---|---|---|---|---|---|---|---|
| | Range | | Peak | Range | | Peak | Range | | Peak |
| | Epiyear/epiweek | Date | Epiyear/ epiweek | Epiyear/epiweek | Date | Epiyear/ epiweek | Epiyear/epiweek | Date | Epiyear/ epiweek |
| **1** | 2020/10–2020/39 | 1 March 2020–6 September 2020 | 2020/28 | 2020/10–2020/40 | 1 March 2020–3 October 2020 | 2020/29 | 2020/10–2020/42 | 01 March 2020 – 17 October 2020 | 2020/29 |
| **2** | 2020/40–2021/13 | 27 September 2020–03 April 2021 | 2021/01 | 2020/41–2021/13 | 4 October 2020–3 April 2021 | 2021/01 | 2020/43–2021/13 | 18 October 2020 – 03 April 2021 | 2021/01 |
| **3** | 2021/14–2021/44 | 4 April 2021–06 November 2021 | 2021/26 | 2021/14–2021/44 | 4 April 2021–6 November 2021 | 2021/27 | 2021/14–2021/46 | 04 April 2021 – 20 November 2021 | 2021/27 |
| **4** | 2021/45–2022/12 | 7 November 2021–26 March 2022 | 2021/49 | 2021/45–2022/13 | 7 November 2021–2 April 2022 | 2021/50 | 2021/47–2022/14 | 21 November 2021 – 09 April 2022 | 2021/52 |
| **5** | 2022/13–2022/32 | 27 March 2022–13 August 2022 | 2022/19 | 2022/14–2022/32 | 3 April 2022–13 August 2022 | 2022/19 | 2022/15–2022/32 | 10 April 2022 – 13 August 2022 | 2022/19 |
